# Urinary stone segmentation: A computationally efficient yet consistently effective approach using image processing

**DOI:** 10.64898/2026.09.16.26357464

**Authors:** Binh D. Le, Seo-Hyeon Yang, Hao T. Nguyen, Kyung-Jin Oh, Ilwoo Park

**Author notes:** These authors have contributed equally to this work and share the corresponding authorship (Kyung-Jin Oh, Ilwoo Park). **Correspondence**: Ilwoo Park, Ph.D., Department of Radiology, Chonnam National University Hospital, 42 Jebong-ro, Dongu, Gwangju, Korea (61469), Kyung-Jin Oh, M.D., Ph.D., Department of Urology, Chonnam National University Hospital, 42 Jebong-ro, Dongu, Gwangju, Korea (61469).

## Abstract

**Objectives:** To assess the effectiveness of our proposed urinary segmentation method based upon image processing, and to contrast its performance against various approaches.

**Subjects and Methods:** Non-contrast-enhanced computed tomography (NCCT) scans of instances with stone disease were collected. An experienced urologist and a senior radiology resident generated ground truths by manually drawing the regions of interest (ROIs) of stone samples. The proposed method took loosely defined ROIs as input and included three steps: (1) finding *mean_HU_rim*, the mean Hounsfield unit (HU) of the stone rim, on each NCCT slice by an edge detection operator; (2) considering pixels with HU higher than the minimum value of all the *mean_HU_rim*; and (3) removing noise. The entire data was divided into a developing set of 406 samples to evaluate the performance of the proposed method as well as to fine-tune a *nnUNetv2* model and a comparison test set of 125 samples to compare the performance and inference time of various segmentation methods. In addition, the reproducibility of our method was tested on the 30 samples randomly selected from the entire data.

**Results:** A total of 531 stone samples from 287 instances were included. Our method showed a high level of agreement with the ground truth in the developing set, with the median (interquartile range) Dice coefficient of 0.86 (0.81 – 0.89). The reproducibility of our method was remarkably high with the median (interquartile range) Dice coefficient of 1.0 (0.991 – 1.0). On the comparison test set, our semi-automatic segmentation outperformed the fixed thresholding and deep-learning-based methods. Lastly, the inference time of the proposed method was less than a second.

**Conclusion:** The proposed semi-automatic approach provided a consistently reliable, simple, and robust method to segment urinary stones.

## 1. INTRODUCTION

Urinary stone disease is among the most prevalent health burdens. The prevalence was estimated at approximately 10% in the US, and from 5% to 19.1% in the “stone belt” in Asia ^1,2^. The incidence is on a rise largely due to the ubiquitous use of imaging modalities ^3^.

Detecting stone on radiographic images plays a crucial role in diagnosis and management. It is evident that radiographic characteristics contribute to the pre-interventional prediction of composition as well as the successful rate of treatment ^4^. Predicted composition could guide the selection of treatment regimens such as medical chemolysis, extracorporeal shock wave lithotripsy (ESWL), and various surgical methods. Since the prediction of stone composition relies on analyzing the imaging characteristics of radiographic data, the precise delineation of the stone for quantitative analyses is of clinical importance. However, manual delineation is not only time consuming but also susceptible to intra- and inter-reader variation.

Automatic segmentation, therefore, could offer an alternative solution. Different threshold values were adopted to isolate urinary stones, for example, 130 HU ^5^, 200 HU ^6,7^, and 250 HU ^8,9^. The most commonly used cut-off value is 130 HU, which originated from a study on detecting coronary calcification ^10^. Because the biochemical composition of urinary stones differs from that of vascular calcified plaques ^11^, thresholding at 130 HU may not be most appropriate for urinary stones. In a study on the quantification of urinary stone volume, applying flexible thresholding values for each stone was suggested rather than applying fixed threshold for all subjects ^12^.

Thanks to the advance in computer vision, the automatic segmentation of urinary stone using artificial intelligence has become a popular method. For example, Mukherjee et al. segmented stones by binarily thresholding the previously isolated kidneys by an U-Net model ^13^. In another attempt, Babajide et al. modified a deep learning algorithm for segmenting stones from 94 patients and reported the Dice coefficient of 0.66 compared to human inputs ^14^. When more complex models were implemented, the Dice coefficient was reported to be as high as 0.88 ^15,16^.

In this study, we aimed at developing a simple yet efficient segmentation approach using image processing based upon an edge detection operator. To the best of our knowledge, the use of this method for segmenting urinary stones remains largely unstudied.

## 2. MATERIALS AND METHODS

### 2.1. Research population

Approval for this retrospective study was obtained from the Institutional Review Board of Chonnam National University Hospital (IRB No. 2022-434), and the requirement for informed consent was waived.

We utilized the dataset from our previous study ^17^. Individual stones from the instances of patients, who were diagnosed with stone disease and managed at the Department of Urology, Chonnam National University Hospital from January 2019 to December 2021, were included as individual samples. Preoperative non-contrast-enhanced computed tomography scans (NCCTs) of the instances were retrieved and screened. In addition to the exclusion criteria from our previous work (listed in the green table in Figure 1), we further excluded samples that possessed closely located stones (listed in the red table in Figure 1) because we aimed at developing a semi-automatic segmentation algorithm for each individual stone.

**Figure 1.**
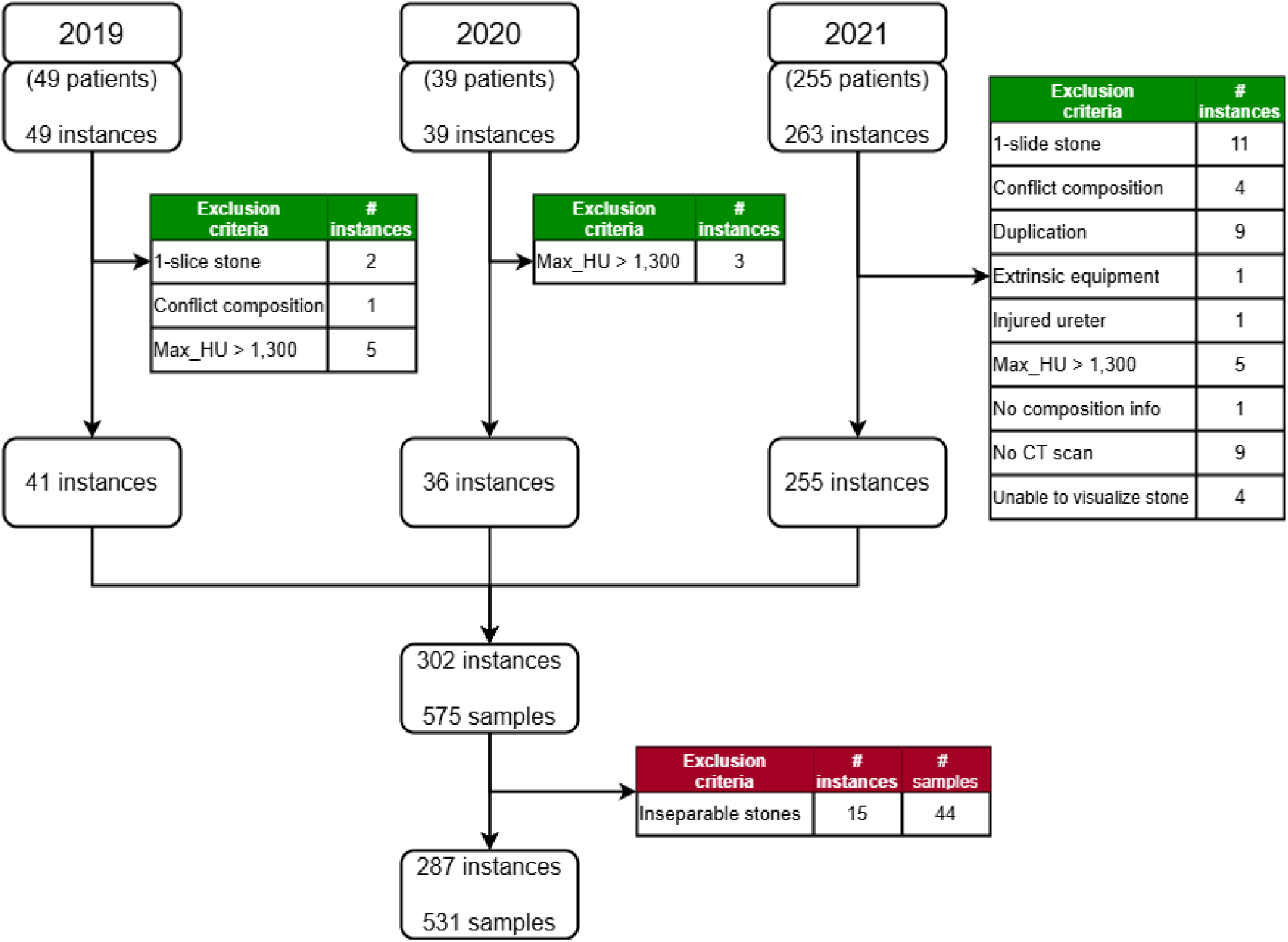
Flowchart of the research scheme. Exclusion criteria in the *green* tables were adopted from the previous, while an additional exclusion criterion in the *red* table was applied to the current study. *Max_HU > 1,300*, pure uric acid-labeled instances with unusually high maximum HU; C*onflict composition*, instances with inconsistent composition analyses; *1-slice stone*, samples visualized only on a single CT slice; *Extrinsic equipment*, the presence of medical instruments; *Injured ureter*, visualization of the stone interfered by an injured ureter; *No composition info*, no entry of stone composition available; *No CT scan*, no CT scan was found; *Unable to visualize stone*, stones were too small to be visualized; *Duplication*, instances that were already included.

### 2.2. Data division

From the total of 531 samples, we defined three data sets: developing, comparison test and reproducibility test sets. The entire samples were randomly divided into the developing and comparison test sets. The developing set included 406 samples that were used to assess the performance of the proposed semi-automatic segmentation approach. This data set was also used for fine-tuning one of the advanced deep learning-based segmentation algorithms against which our approach was compared. The comparison test set consisted of 125 samples that were utilized for comparing the performance and inference time of our method with those of different segmentation methods. Lastly, the reproducibility test set consisted of 30 samples that were randomly selected from the entire data to investigate the reproducibility of the proposed method (Figure 2a).

**Figure 2.**
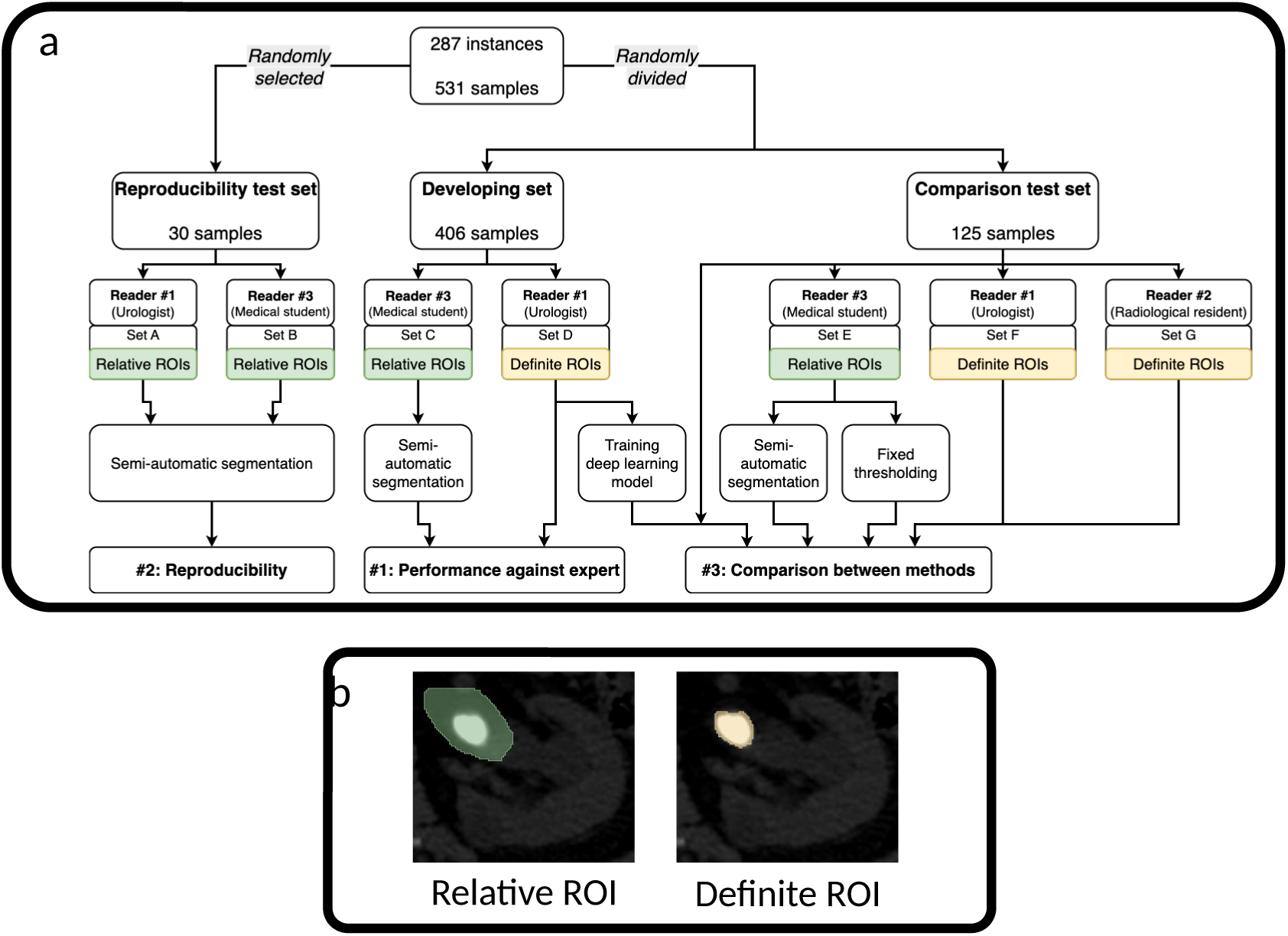
Data division and region of interest definition. *a,* Diagram of data division and arrangement. Three readers were involved in generating the relative and definite regions of interest (ROIs). These ROIs and the corresponding non-contrast-enhanced computed tomography scans were adopted for three evaluation tasks. The first was to evaluate the performance of the semi-automatic segmentation algorithm against an expert (Reader 1) using set C and D. The second task was to assess the reproducibility of the semi-auto segmentation method using set A and B. The last task was to compare the segmentation effectiveness of different methods using the set D, E, F, and G. For this task, a deep learning model (*nnUNetv2*) trained on set D was executed to predict ROIs of the samples in the comparison test set, while the semi-automatic segmentation and fixed thresholding methods took the set E as input. All these segmentation approaches were compared to definite ROIs from Reader 1 (set F) and Reader 2 (set G). *b,* An example of a pair of relative and definite ROIs.

### 2.3. Region of interest definition

On the NCCTs, the delineation process were conducted in a magnified “bone window” setting of an open-source software, *3D Slicer v5.2.2* ^18^. Samples were manually delineated by three independent readers including a urologist with 4 years of experience (Reader 1), a senior radiology resident (Reader 2), and a junior medical student (Reader 3). The definite regions of interest (ROIs) of stones that were drawn by Reader 1 and Reader 2 were considered as ground truth. Reader 1 was responsible for the definite ROIs of all 531 samples; meanwhile, Reader 2 drew the definite ROIs of 125 samples for the comparison test set (Figure 2a).

To simulate clinical scenario where the segmentation can be performed effortlessly without the time-consuming process of stone delineation, relatively loosely defined ROIs (relative ROIs, hereafter) encompassing both the stone of interest and the large surrounding area were generated and used as inputs to the semi-automatic segmentation algorithm. Reader 3 drew the relative ROIs for all 531 samples (set C and E, Figure 2a). For the reproducibility test set, 30 samples were randomly chosen to generate the sets of relative ROIs by Reader 1(set A, Figure 2a) and Reader 3 (set B, Figure 2a) separately. Relative ROIs served as inputs to either the semi-automatic segmentation algorithm or the fixed thresholding methods. Figure 2b illustrates an example of a pair of relative and definite ROIs.

### 2.4. Semi-automatic segmentation algorithm

The development of our approach was based on the observation of urinary stone position and formation. Firstly, the attenuation of stone was always significantly higher than its surrounding soft tissue or fluid; therefore, the degree of HU gradient between the rim of stone and its surrounding should be the highest, which led us to use an edge detection operator. Secondly, the HU value of the rim was normally lower than that of the core due to the difference in crystal density. Thus, the pixels with the HU values higher than the HU of rim should represent the stone. A pseudocode of our algorithm for the semi-automatic segmentation of each stone sample is presented in Figure S1 and graphically illustrated in Figure S2.

### 2.5. Performance evaluation

We evaluated the performance of the proposed method via three tasks. Firstly, the performance of segmentation was assessed using the developing set by calculating Dice coefficient between the result of semi-automatic segmentation (set C, Figure 2a) and the definite ROIs from Reader 1 (set D, Figure 2a). Secondly, the reproducibility of our method was assessed using the 30 samples from the reproducibility test set. Reader 1 (set A, Figure 2a) and Reader 3 (set B, Figure 2a) generated relative ROIs separately and their outputs from the proposed algorithm were compared. Lastly, the performance of our proposed method was compared with those of the fixed thresholding and deep learning methods using the comparison test set.

For the fixed thresholding approach, we adopted commonly used cut-off values, including 130 HU ^5^, 200 HU ^6,7^, and 250 HU ^8,9^. For the deep learning segmentation approach, we utilized the *nnUNetv2* model. This is an update from the original architecture, *nnUNet*, which is well known for semantic segmentation in the biomedical domain ^19^. It demonstrated notable achievements in multiple computer vision challenges ^20^ and has continuously served as a baseline and a framework for method development ^21,22^. In this study, we fine-tuned the *nnUNetv2* model with the developing set (set D, Figure 2a) and executed inference on the comparison test set. The configurations for training the *nnUNetv2* model were set as default for high 3D image resolution (*3d_fullres*) except that 250 training epochs and no mirroring augmentation were used. The results from the proposed, fixed thresholding, and deep learning approaches were compared to the definite ROIs from Reader 1 (set F, Figure 2a) and Reader 2 (set G, Figure 2a). In addition, inference time were measured and compared between the proposed and deep learning methods.

All calculations and algorithms were implemented in Python 3.11. Experiments were run on a computer with an *Intel* Core i7-11700K CPU at 3.6GHz, an *NVIDIA* GeForce RTX 3060 (12GB) GPU using 64 GB DDR4 of RAM, and two *Samsung* 980 PRO 1TB SSDs, running on Windows 11 operating system version 24H2.

## 3. RESULTS

### 3.1. Data overview

The age of patients who were included in 287 instances was 60.3 ± 17.5 years (mean ± standard deviation) with the male/female ratio of 2.1. NCCTs of 193 instances were acquired from our institution, while those of 94 instances were acquired from 38 outside hospitals. A total of twenty-four scan systems were used with various CT settings. The details of scan settings are listed in Table S1.

### 3.2. Performance of the semi-automatic segmentation compared to the expert

The results from the semi-automatic segmentation demonstrated a high level of agreement with Reader 1 on the developing set. The median (interquartile range) Dice coefficient between the ROIs from Reader 1 and semi-automatic segmentation were 0.86 (0.81 – 0.89) (Figure S3).

### 3.3. The reproducibility of the semi-automatic segmentation method

The proposed method showed a high degree of reproducibility. When provided with two sets of relative ROIs that were created by different readers (a urologist and a medical student), the semi-automatic segmentation algorithm generated identical ROIs in most cases with the median (interquartile range) Dice coefficient of 1.0 (0.991 – 1.0).

The minimum Dice coefficient among the 30 pairs of semi-automatically segmented ROIs was 0.96 (Figure S4). Illustrative examples demonstrating the reproducibility of the proposed segmentation method are presented in Figure S5.

### 3.4. Comparison of the semi-automatic segmentation with other methods

The proposed semi-automatic segmentation approach showed a superior performance over the fixed thresholding as well as the deep learning methods. When the outputs of the segmentation were compared with the definite ROIs drawn by the two experts on the comparison test set, the proposed method achieved higher Dice coefficients than the other methods. The mean ± standard deviation (SD) Dice coefficients of the semi-automatic segmentation in comparison to Reader 1 and Reader 2 were 0.85 ± 0.07 and 0.85 ± 0.05, respectively. In contrast, the mean ± SD Dice coefficients of the 130-HU-fixed thresholding method in comparison to Reader 1 and Reader 2 were 0.73 ± 0.13 and 0.84 ± 0.11, respectively, while the same metrics for the deep learning model were 0.45 ± 0.35 and 0.42 ± 0.32, respectively. The Dice coefficients of the proposed method were significantly higher than those of other segmentation methods for both Reader 1 and Reader 2 (p < 0.05 by paired T-test), except for the 130-HU-fixed thresholding method for Reader 2, whose performance was comparable to ours (p = 0.14 by paired T-test, refer to Table S2 for more details).

In contrast to the thresholding and deep learning methods, the proposed method demonstrated consistent results when the outputs were compared to different ground truths (Figure 3a). When the two Dice coefficients measured by the ground truths from the two experts were compared to each other using the paired T-test, only the semi-automatic segmentation method demonstrated a null hypothesis, meaning that there was no significant difference between the two Dice coefficients (p = 0.32, Table S2). The two Dice coefficients for all the other segmentation methods were significantly different between the two readers (p < 0.001, Table S2).

**Figure 3.** Comparison of performance between various segmentation methods on the comparison test set. *a*, Dice coefficients and the corresponding box and violin plots obtained by comparing the segmentation outputs with the two ground truths (Reader 1 and Reader 2). *b,* A scatter graph showing the inference time of the proposed and deep learning segmentation methods plotted over the number of slices in input images. Each slice was a 2-dimensional image with 512×512 pixels. *Semi-auto,* the proposed semi-automatic segmentation method; *130HU, 200HU, and 250HU,* fixed thresholding at 130 HU, 200 HU, and 250 HU, respectively; *nnUNetv2,* the nnUNetv2 model fine-tuned on the developing set; *nnUNetv2_withGPU* and *nnUNetv2_noGPU,* the fine-tuned nnUNetv2 model with and without the use of GPU, respectively.

The inference time of the *nnUNetv2* model was proportional to the number of NCCT slices of the input. When utilizing GPU for inference, the model took 10 - 27 seconds for each sample with all NCCT slices provided as input. The inference time rose remarkably to 6.4 - 17.6 minutes without GPU. When the input was intentionally limited to only NCCT slices of interest that showed the stones, the inference time of the fine-tuned *nnUNetv2* model decreased, but was still significantly longer (0.5 – 13.1 seconds with GPU and 13 - 493 seconds without GPU) than that of our method (0.05 – 0.39 seconds). In contrast, the proposed semi-automatic segmentation provided the outputs in less than a second without the use of GPU regardless of the number of input slices (Figure 3b).

## 4. DISCUSSION

Using a simple and cost-effective approach, we demonstrated the semi-automatic segmentation of urinary stones with a high accuracy and robustness. When the outputs from our method were compared to the ground truth in the developing and comparison test sets, we consistently found the median Dice coefficients of 0.86 to 0.87 with the interquartile ranges within 0.80 – 0.90. These findings suggest that our approach consistently produced the effective segmentation outputs that exhibited good agreements with experts. In addition, the proposed method showed a high level of reproducibility regardless of whether the input (relative ROI) was created by an expert or not. This implies that our method does not require a specialist’s input but rather can be reliably executed by general healthcare workers.

Compared to the previous studies which utilized more sophisticated approaches, the proposed semi-automatic segmentation was able to provide comparable or even superior performance. For example, Kim et al. developed three separate deep learning models for axial, coronal, and sagittal images and integrated them to achieve the Dice coefficient of 0.88 ^16^. Li et al. trained five deep learning algorithms, SegNet, DeepLabV3+, 3D U-Net, UNETR, and Res U-Net, in two stages and reported that Res U-Net achieved the best mean Dice coefficient of 0.81 ^15^. Another effort by Babajide et al. introduced a modified deep learning model and reported a Dice coefficient of 0.66 ^14^.

Although these comparisons can provide useful information regarding the capability of our method, the evaluation and comparison of various methods using different test data require cautious approach and could lead to a biased interpretation. To overcome this issue, we utilized a separate data set, the comparison test set, to compare the performance of our proposed semi-auto segmentation with those of fixed thresholding at various cut-off values and deep-learning-based-segmentation approaches. The output of stone segmentation using the thresholding at fixed HU values depends on the choice of cut-off value since applying a high cut-off value helps in isolating the core but misses out the outer part or boundary of urinary stones. As a result, the higher the applied cut-off values were, the lower the value of Dice coefficient became when this approach was tested on the ground truth provided by experts (Figure 3a). A more flexible approach, such as the method proposed by Demehri et al., has also been considered ^12^. The authors segmented stones by thresholding at the half of the measured mean attenuation of each stone. However, this kind of thresholding method requires a thoughtful consideration and effortful delineation of the area to measure HU, which is likely to be subjective and inconsistent. In contrast, we proposed a flexible and simple approach, based upon the nature of the attenuation gradient between the stone and its surrounding area, which required a minimal human effort. Our proposed method demonstrated higher Dice coefficients than the fixed thresholding across various common cut-offs on the comparison test set, which suggests that our approach may provide an alternative to overcome the limitations of the fixed HU thresholding method.

We further assessed the performance of a well-known deep learning framework for biomedical segmentation, *nnUNetv2,* in a direct comparison to our method on the same comparison data set. Dice coefficient and inference time were assessed and compared between the two methods. We found that the fine-tuned *nnUNetv2* model displayed a much lower performance in capturing the stone of interest defined by the two experts than our proposed semi-automatic segmentation (Figure 3a). The suboptimal output from the *nnUNetv2* model may stem from the fact that, unlike solid organs, urinary stones are not stationary in spatial position. The positional variability of stone occurrence was a significant challenge for the deep learning model if no guidance was provided in terms of location. The guidance can be in the form of isolating the kidney with stones by another deep learning model ^13,15^, or by a series of preprocessing steps involving a kidney localization ^23^. Of note, these attempts only focused on urinary stones in kidney and did not include stones in ureter or bladder. In many cases of our current work, the fine-tuned *nnUNetv2* model either rendered a wrong prediction of other structures (e.g., skeleton bones and calcification) as urinary stones or failed to identify the stone of interest. These false positive predictions often occurred when the entire abdominal slices of NCCT scan were used as input (Figure S6). One of the potential solutions to overcome this limitation is using semantic segmentation models which label and isolate the collecting system prior to the segmentation of stones.

Another solution may be obtained by deliberately selecting and limiting the NCCT slices that contain the stone of interest. We tried this solution on the fine-tuned *nnUNetv2* model using the comparison test set; however, it still generated wrong predictions in several cases (see Figure S7 for an example). In short, utilizing deep learning-based models for the task of urinary stone segmentation still needs a human monitor and correction. On the contrary, our proposed method requires a minimal input from human once at the beginning, regardless of the number of CT slices provided.

Another advantage of the proposed semi-automatic segmentation method over the deep learning approach was that it required significantly less computational resources. We investigated the inference time of our method and compared it with those of the fine-tuned *nnUNetv2* models with and without the use of GPU (Figure 3b). The results showed that our approach, which used a moderate computational power relying on CPU only, was able to provide the output with a high level of accuracy in just less than a second. Meanwhile, the fine-tuned *nnUNetv2* required significantly longer time of inference even if a larger computation power replying on GPU was used. This implies that our approach can be readily integrated into other frameworks and is ready to be executed in various system configurations for the task of urinary stones segmentation.

One of the interesting findings in this study came from comparing the distributions of the Dice coefficients measured by the ground truths from two different readers. In the box and violin plots of the two Dice coefficients from Reader 1 and 2 (Figure 3a), it appeared that the semi-automatic segmentation method exhibited relatively similar distributions between the two Dice coefficients, while in other methods, the distributions between the two Dice coefficients appeared very dissimilar. When the distributions between the two Dice coefficients were quantitatively analyzed, we found that the Dice coefficient calculated by the ground truth from Reader 1 was not significantly different from that from Reader 2 in only the proposed method (p = 0.32 by paired T-test). In all the other methods, the two Dice coefficients were significantly different (Table S2). It is worth noting that Reader 2, who was a senior radiology resident, created the ground truth solely based on the radiographic finding, which led to a relatively conservative approach in delineating stones: only bright pixels corresponding to the stone core were included while the grey pixels in the boundary were left out. In contrast, Reader 1, who was a urologist with 4 years of experience, included more grey pixels in the stone boundary because stone formation is a continuous process from the core to the surface and the grey zone on the stone surface can contain important information about the composition of stone. Regardless of these differences in the ground truths from two experts, our segmentation method generated consistent outputs with a high accuracy. In addition, the proposed model demonstrated consistently high performances across 125 samples in the comparison test set that were tested, while other methods showed widely varied performances among the tested samples (Figure 3a). For example, the mean Dice coefficient of our method from Reader 2 was 0.85 with the small SD of 0.05 (Table S2). Although the fixed thresholding method at 130 HU demonstrated the comparable mean Dice coefficient of 0.84 from Reader 2, its SD was 0.11, indicating a relatively wider range of Dice coefficient.

One of the limitations of the current work is that the segmentation process was not fully automated and the generation of human-defined relative ROIs was still required for input. However, the deep learning model intended for full automation, such as *nnUNetv2*, is likely to need a human monitor and correction to achieve optimal results. The proposed method was faster, more consistently reliable, and computationally efficient than *nnUNetv2*. Future works may focus on adopting the semantic segmentation method to separate the collecting system prior to the stone segmentation, which may minimize human interference.

In summary, our results suggest that the proposed semi-automatic approach provided a consistently reliable, simple, and robust way to segment the urinary stone. Due to its relatively low demand for computational power, this method can be readily integrated into other frameworks or systems.

## ABBREVIATIONS

CT: Computed tomography
HU: Hounsfield unit
NCCT: Non-contrast-enhanced computed tomography
ROI: Region of interest
SD: Standard deviation

## ACKNOWLEDGEMENTS

This research was funded by Institute of Information & communications Technology Planning & Evaluation (IITP) under the Artificial Intelligence Convergence Innovation Human Resources Development (IITP-2023-RS-2023-00256629) funded by the Korea government (MSIT), the National Research Foundation of Korea (NRF) funded by the Ministry of Education (RS-2025-25430545), Chonnam National University (2025-0384-01), and Chonnam National University Hospital Biomedical Research Institute (BCRI25010).

## DATA AVAILABILITY

The imaging data used in this study are not publicly available owing to institutional restrictions imposed to protect patient privacy. The source code is not publicly available; further inquiries may be directed to the corresponding author.

